# Variation in COVID-19 excess mortality by age, sex, and province within Italy

**DOI:** 10.1101/2021.07.14.21260494

**Authors:** Nathaniel Henry, Ahmed Elagali, Michele Nguyen, Michael Chipeta, Catrin Moore

## Abstract

Although previous evidence suggests that the infection fatality rate from COVID-19 varies by age and sex, and that transmission intensity varies geographically within countries, no study has yet explored the age-sex-space distribution of excess mortality associated with the COVID pandemic. By applying the principles of small-area estimation to existing models formulations for excess mortality, this study develops a method for assessing excess mortality across small populations and assesses the pattern of COVID excess mortality by province, year, week, age group, and sex in Italy from March through May 2020. We estimate that 53,200 excess deaths occurred across Italy during this time period, compared to just 35,500 deaths where COVID-19 was registered as the underlying cause of death. Out of the total excess mortality burden, 97% of excess deaths occurred among adults over age 60, and 68% of excess deaths were concentrated among adults over age 80. The burden of excess mortality was unevenly distributed across the country, with just three of Italy’s 107 provinces accounting for 32% of all excess mortality. This method for estimating excess mortality can be adapted to other countries where COVID-19 diagnostic capacity is still insufficient, and could be incorporated into public health rapid response systems.

## 2 Introduction

Italy received international attention as one of the first countries outside of China to experience a major COVID-19 outbreak. On March 9, 2020, Italy announced a nationwide lockdown to stem community transmission, and deaths peaked two weeks later during the week of March 25^1^. After that peak, deaths declined for the following two months, and lockdown restrictions were gradually eased starting in late May^2^. In total, between February and August 2020, approximately 35,500 COVID-19 deaths were registered across Italy, equivalent to approximately 60 deaths per 100,000 people^3^.

Aside from the timing and magnitude of the first wave of COVID-19 transmission within Italy, two notable features set its COVID-19 epidemic apart from those in other European countries. First, registered COVID-19 cases and deaths were unevenly distributed across the regions of Italy^4^, and that cause-specific mortality from COVID-19 varied by age and sex^5^. Registered COVID-19 deaths were highest in the northern regions of the country, particularly in the region of Lombardy, where the registered COVID-19 death total amounted to over 160 deaths per 100,000 population (Figure 1). Another salient feature of the Italian epidemic was the early recognition among health authorities that registered COVID-19 deaths under-counted the full mortality burden of the epidemic. In May 2020, the Italian National Institute of Statistics (Istat) reported that while 13,700 COVID-19 deaths had been registered across Italy between 20 February and 31 March, deaths from any cause had increased by 25,300 compared to an expected baseline during the same time period, suggesting that the full mortality burden of the COVID-19 epidemic was nearly double what had been previously reported^6^.

**Figure 1:**
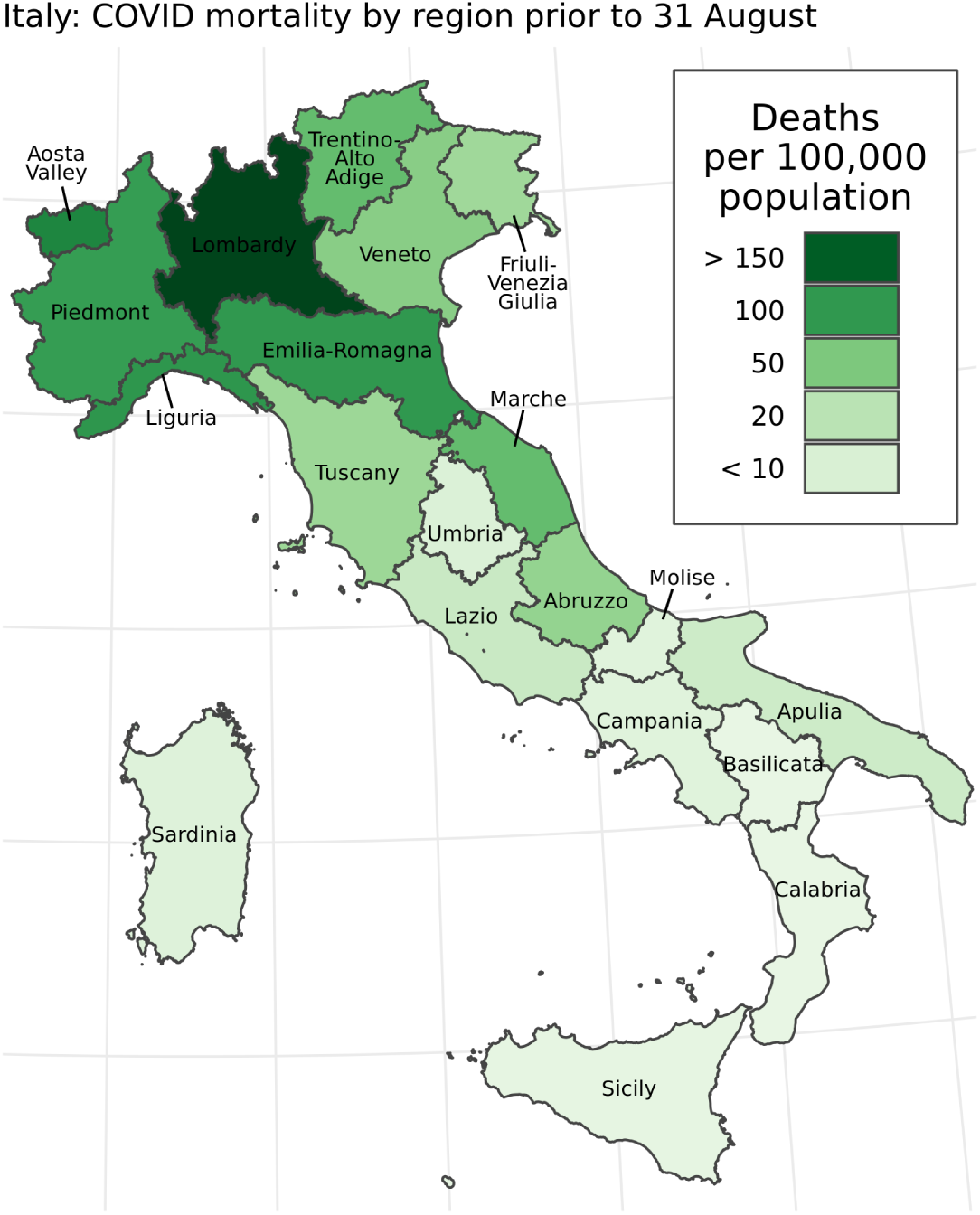
Registered COVID-19 deaths per 100,000 population by Italian region prior to August 31, 2020. Nationally, 35,500 deaths were registered during this time, or approximately 59 deaths per 100,000 population.

Istat and other research groups drew conclusions about the mortality burden of the COVID-19 pandemic based on a series of excess mortality analyses. Excess mortality analyses attempt to measure the net effect of a discontinuity, such as a COVID-19 outbreak, on all-cause mortality. This is a two-step process: first, the investigator constructs an estimated baseline number of deaths expected during the period in question. While various methods have been used in the past to construct this baseline^7–9^, previous studies in Italy have averaged the number of deaths recorded by week in the years from 2015 through 2019 to generate a baseline estimate for the same weeks in 2020^2,10^. Next, the investigator compares that expected baseline with the count of recorded deaths during the same time period. Excess deaths, a count, are measured as the difference between the observed death count and the expected baseline. Standardized mortality ratios (SMRs) are measured as the ratio between the observed death count and the expected baseline^11,12^. Excess mortality analyses of the COVID-19 pandemic are becoming widely used in the media: among others, the Economist, the Financial Times and the New York Times have estimated excess mortality across dozens of countries in Europe, the Americas, and Asia^13–15^. Previous investigations have also examined how excess mortality analysis can capture deaths caused by COVID-19 but attributed to other causes, as well the indirect mortality burden of the COVID-19 pandemic^8,16^.

This study explores another central issue for excess mortality estimation: how can we detect increases in mortality at the local level or across multiple age groups, where the expected number of baseline deaths in each subpopulation of interest is relatively low? This question addresses a tension central to many forms of public health surveillance, where the imperative to identify clustering and high-risk subgroups in health surveillance data must be balanced against reduced study power and possible biases associated with small samples^17^. Any suitable approach for estimating small-group excess mortality must quantify uncertainty due to stochastic variation as well as limited data informing the baseline. Even at the national level, weekly estimates of COVID-19 excess mortality presented without uncertainty intervals can leave viewers confused about what constitutes a meaningful departure from the baseline.

To better understand the relationship between registered COVID-19 deaths and excess mortality across Italy, we developed a model to estimate excess mortality by age group, sex, and week across the country’s 107 provinces. Our approach estimated counterfactual baseline mortality rates for each of these groups from March through August 2021, based on mortality rates and predictive covariates observed from January 2015 through February 2021. Our baseline mortality model combined elements from a widely-used Poisson generalized linear modeling (GLM) strategy for mortality estimation^9^; a structured province-year-age random effect that draws power from local correlations in mortality across those three dimensions, based on disease mapping principles^18^; and a Fourier curve-fitting method to fit capture seasonal trends in age-specific all-cause mortality^7^. Once baseline mortality was estimated for each week and subpopulation in the study period, we compared these estimates to observed counts from vital records over the same period while preserving uncertainty in the expected baseline mortality. Here, we present major findings on excess deaths (the difference between observed and baseline death counts) as well as SMRs (the ratio between observed and expected deaths) across these subpopulations of Italy.

## 3 Results

### 3.1 Excess mortality exceeds registered COVID-19 deaths

At the national level, the aggregated results from our model generally agreed with previous national studies on the timing and magnitude of excess deaths associated with the COVID-19 pandemic across Italy. Figure 2, below, shows the estimated weekly death count (in brown) as well as the observed weekly death count (in black) from 26 February through 31 August 2020 across Italy. In line with other studies, the model results suggest that excess mortality peaked on the week of March 25, on the same week as the peak in registered COVID-19 deaths, and then consistently fell until approximately returning to baseline by the end of May 2020, coinciding with the lifting of most lockdown measures across Italy^2^. At no point between June and August 2020 did the observed death count exceed the upper bound of the 95% uncertainty interval for baseline deaths: as a result, the remainder of this section will focus on excess mortality during the 13-week period from 26 February through 26 May 2020. During these 13 weeks, we estimate that 53,200 excess deaths (95% uncertainty interval 26,500 to 79,700) were associated with the COVID-19 pandemic, compared with 35,500 deaths registered with COVID-19 as the underlying cause during the same period.

**Figure 2:**
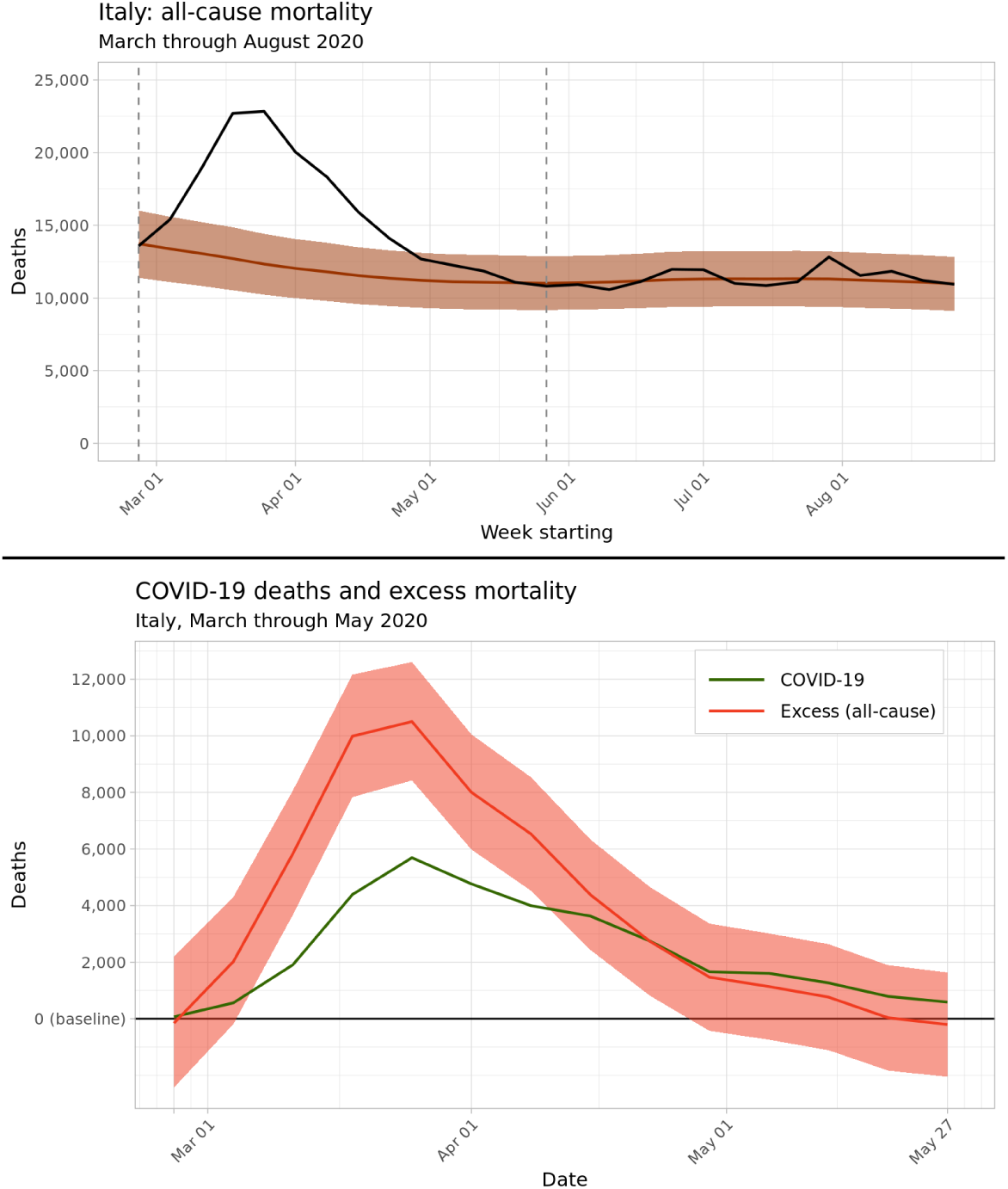
*Top* Predicted baseline and observed deaths aggregated nationwide across Italy, March through August 2020. Bounds of the 95% uncertainty interval for predicted baseline deaths are shaded in light brown. *Bottom* Estimated weekly excess deaths across Italy from March through May 2020, in red, compared with medically-certified deaths from COVID-19, in green.

### 3.2 Excess deaths and sex

Our findings can be aggregated across sexes and provinces to understand the age structure of excess mortality across Italy. Figure 3 plots weekly excess deaths across the five modeled age categories, while Table 1 lists estimated excess deaths by age and sex grouping at the national level. Excess deaths were overwhelmingly concentrated in older age groups: of the estimated 53,200 excess deaths in Italy from March through May 2020, an estimated 51,600, or 97.0%, of these excess deaths occurred among adults aged 60 and above, while 36,400, or 68%, occurred in adults older than age 80. Among adults aged 90 and above, women experienced 11,000 (5,800 to 16,400) excess deaths, more than double the 4,400 (2,000 to 6,900) excess deaths among men in the same age group; this reflects the sex composition of the oldest age group, where women made up 72.6% of the over-90 population in January 2020.

**Table 1:** Estimated excess mortality by age and sex grouping, aggregated to the national level for the weeks of 26 February through 26 May 2020. Nationally, all age-sex groupings experienced significantly elevated mortality compared to the expected baseline during this period.

| Sex | Age | Excess deaths<br>(95% UI) | Standardized<br>Mortality Ratio<br>(95% UI) | Population on<br>1 Jan 2020,<br>millions (% total) |
| --- | --- | --- | --- | --- |
| Males | 0-59 | 1,081 (918 to 1,240) | 1.16 (1.13 to 1.19) | 21.37 (35.5%) |
| Females | 0-59 | 517 (401 to 627) | 1.13 (1.1 to 1.16) | 21.00 (34.9%) |
| Males | 60-69 | 2,600 (1,152 to 3,990) | 1.29 (1.11 to 1.52) | 3.55 (5.9%) |
| Females | 60-69 | 947 (109 to 1,750) | 1.19 (1.02 to 1.41) | 3.87 (6.4%) |
| Males | 70-79 | 7,703 (4,639 to 10,730) | 1.42 (1.22 to 1.71) | 2.75 (4.6%) |
| Females | 70-79 | 3,991 (1,885 to 6,048) | 1.33 (1.13 to 1.6) | 3.25 (5.4%) |
| Males | 80-89 | 10,649 (4,574 to 16,946) | 1.36 (1.13 to 1.72) | 1.45 (2.4%) |
| Females | 80-89 | 10,308 (3,675 to 17,110) | 1.32 (1.09 to 1.67) | 2.20 (3.7%) |
| Males | 90 | 4,405 (1,979 to 6,879) | 1.36 (1.14 to 1.71) | 0.22 (0.4%) |
| Females | 90 | 11,043 (5,784 to 16,432) | 1.41 (1.18 to 1.76) | 0.58 (1.0%) |

**Figure 3:**
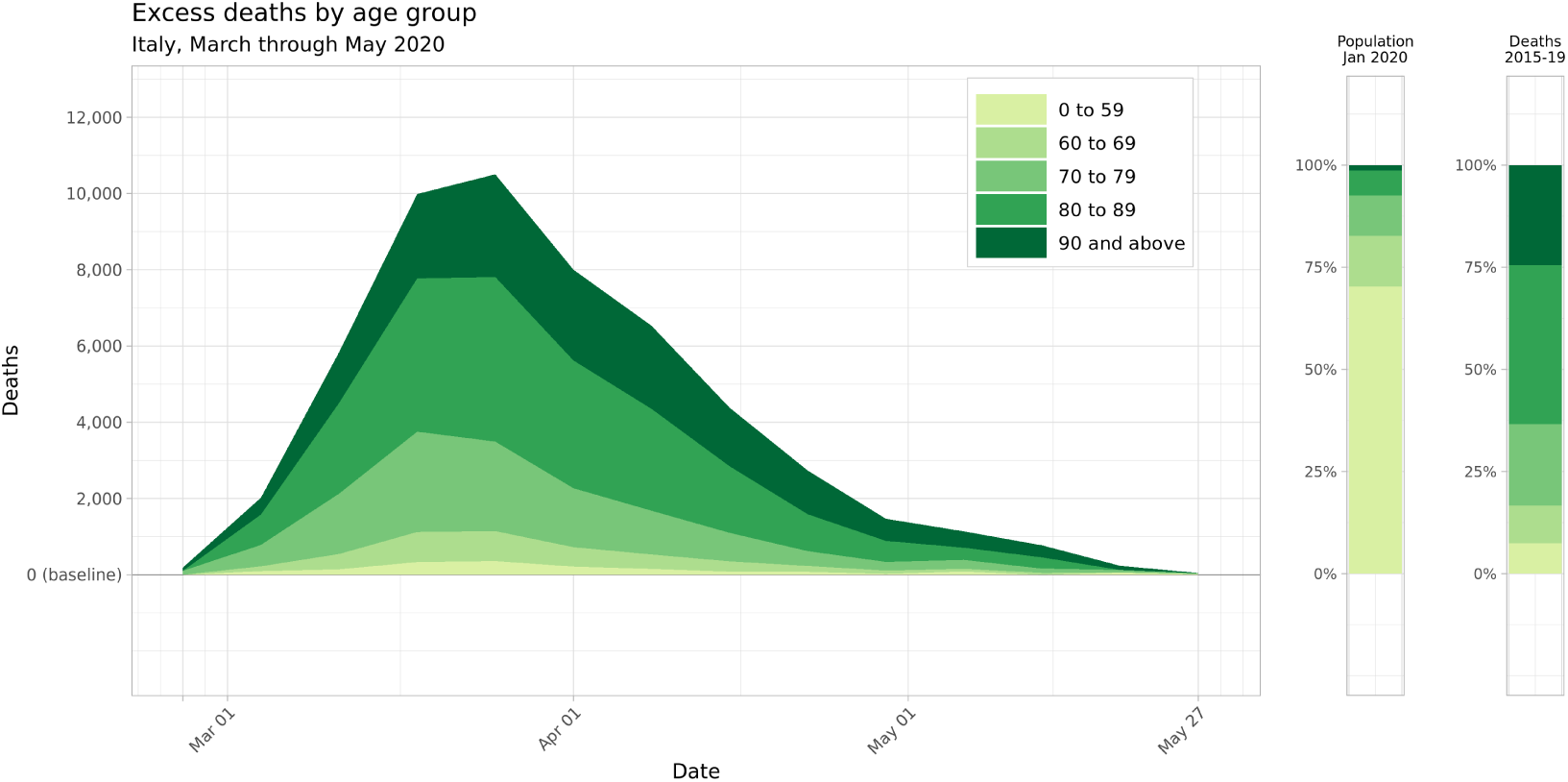
Mean estimates for weekly excess deaths across Italy by age group, March through May 2020. Two nationwide age group breakdowns are shown to the right for comparison: the national age distribution as of 1 January 2020, and the proportion of nationwide deaths that occurred in each age group during the years 2015 through 2019.

The outsized proportion of excess deaths observed within older age categories reflects two aspects of age-structured mortality across Italy during this period: baseline mortality is highest in older age groups even in normal years, and these age groups also experienced a larger relative increase in mortality during the study period. Figure 4 maps the standardized mortality ratio, or the ratio between observed and baseline mortality rates, by age group and province across the entire 13-week period from 26 February through 26 May 2020. Note that the standardized mortality ratio uniformly increases across the northern provinces of Italy when moving from the 0-59 age group to the 60-69 and 70-79 age groups, and remain heightened in the oldest age groups, with some provinces in Lombardy and Emilia Romagna experiencing over three times the expected baseline mortality in older age groups. The greater increase in excess mortality among older age groups can also be expressed by comparing the proportion of excess mortality with each age group with that age group’s share of all-cause mortality in the years 2015 through 2019. Adults over age 60 accounted for 92.6% of all deaths in the years 2015-2019 (2.99 million deaths out of 3.23 million total), but 97.0% of all excess mortality in March through May of 2020 (51,600 of 53,200 excess deaths). Similarly, adults over age 80 experienced 63.4% of all deaths in previous years (2.04 million deaths out of 3.23 million), but 68.4% of excess mortality (36,400 of 53,200 excess deaths).

**Figure 4:**
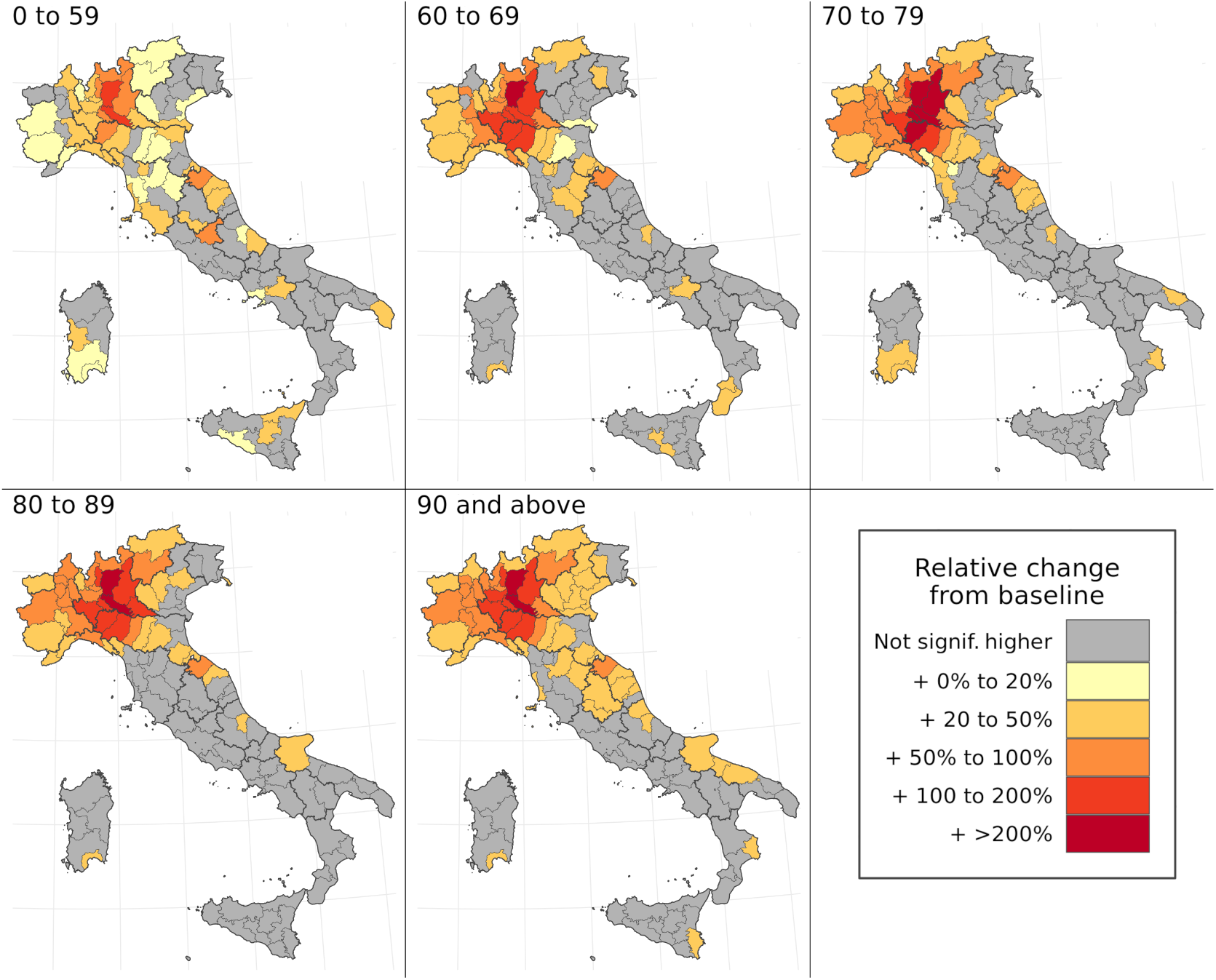
Map of standardized mortality ratios by age group and province over the entire period of March through May 2020. In these figures, significance is defined as falling outside of the 95% uncertainty interval for baseline mortality for a given age group and province during this period.

### 3.3 Spatial concentration

This model is also able to characterize spatial variation in excess mortality during the first wave of the COVID-19 epidemic in Italy. From March through May 2020, just three provinces—Milan, Bergamo, and Brescia, all in the Lombardy region—accounted for 32% of all excess deaths in Italy, with just 9% of the country’s total population. By including just four other provinces across the Piedmont, Emilia-Romagna, and Liguria regions in northern Italy, nearly one-half of all excess deaths are captured in a region that makes up just 16% of Italy’s population. Expanding further, the 22 provinces with the highest number of excess deaths account for three-quarters of all excess mortality, but just 30% of Italy’s national population. Figure 5 shows the locations of these province groupings and each group’s marginal contribution to the total excess mortality curve from March through May 2020.

**Figure 5:**
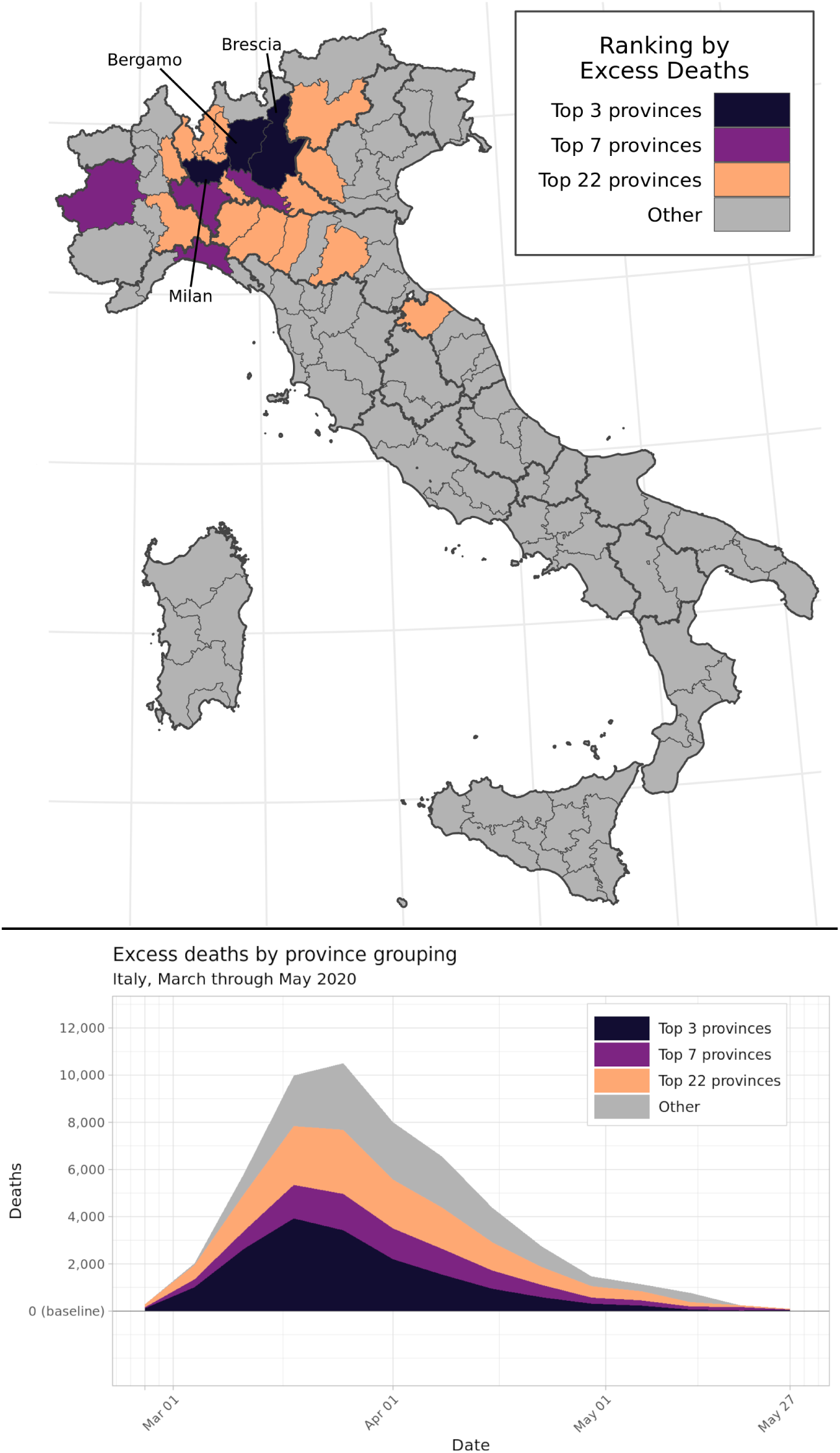
Concentration of excess mortality burden among the 3, 7, and 22 provinces with the greatest number of excess deaths, out of 107 provinces total. These province groupings respectively accounted for over 25%, 50%, and 75% of the national excess mortality burden between March and May 2020. The graph on the bottom left displays mean estimated excess deaths by province grouping and week during this time period.

### 3.4 Diverse experiences of the first wave

Comparing Figures 1, 4, and 5, both the timing and geographical distribution of excess mortality during March through May 2020 seem to follow the time pattern of registered COVID-19 mortality by region in that same period: both peaked during the week of 25 March and were concentrated in the northern regions of the country, particularly Lombardy. Figure 6 complicates this analysis by demonstrating two features of excess mortality that are apparent at the province level, but not the regional or national levels. The first feature identifies the geographic center of the peak excess mortality during the COVID-19 first wave. While most news outlets and previous studies have identified Lombardy region as the center of the COVID-19 outbreak, the top-right panel of Figure 6 suggests that the mortality rate increased most in provinces on the border between southwest Lombardy and northwest Emilia-Romagna, adjacent to the Piedmont and Liguria regions. This border effect suggests that the 107 provinces of Italy may be a more informative unit of analysis for both registered COVID-19 deaths and excess mortality than the country’s 20 regions.

**Figure 6:**
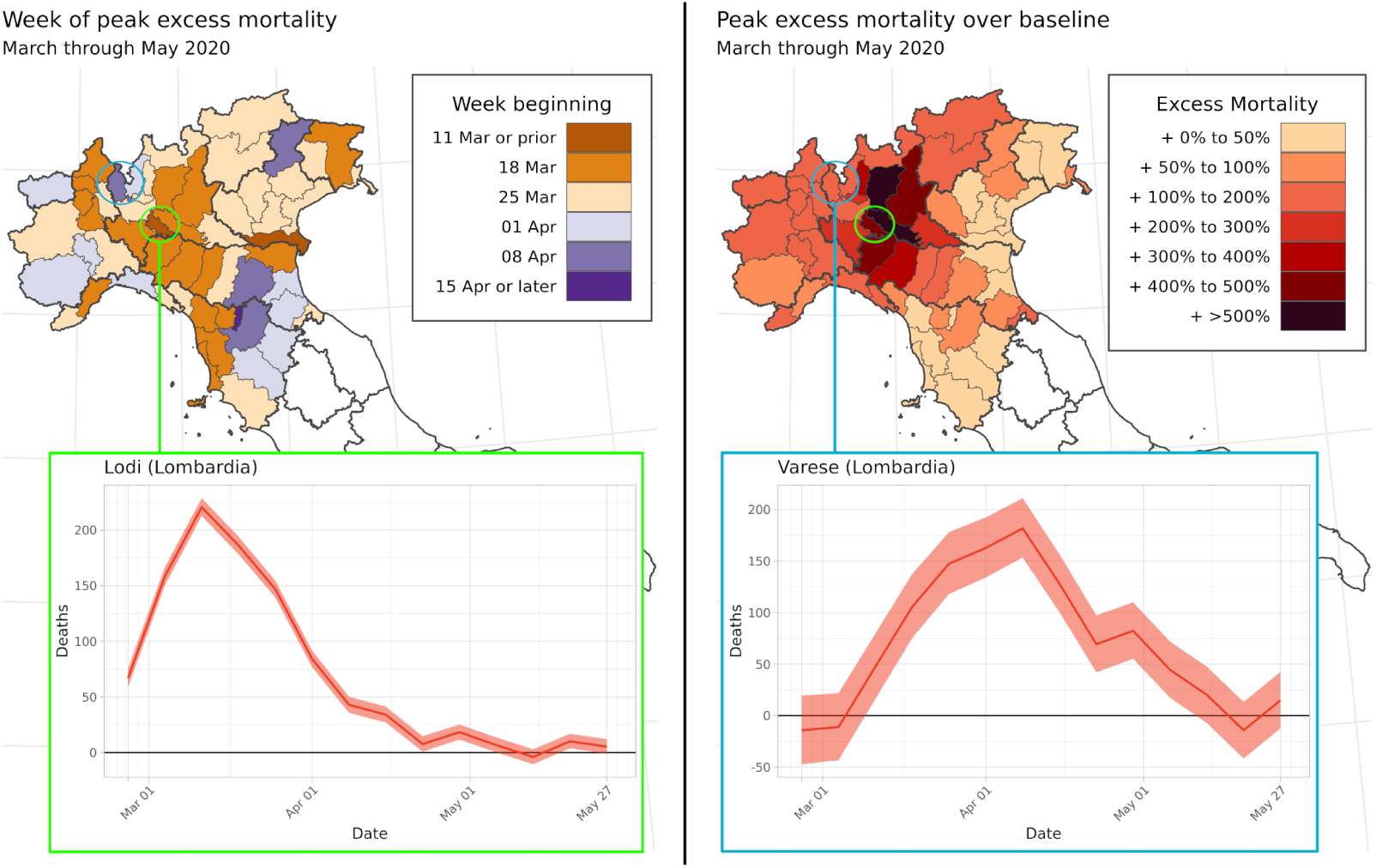
Excess deaths vary across provinces within the Lombardy region. *Top left* Map of Northern Italy showing the week when province’s excess mortality was estimated to peak during the first wave of COVID-19 in March through May 2020. *Top right* Map of the peak weekly standardized mortality ratio (SMR) for each province across Northern Italy across all weeks in March through May 2020. *Bottom left* Estimated excess deaths by week for Lodi province. *Bottom right* Estimated excess deaths by week for Varese province.

A province-level analysis also demonstrates that within high-burden regions, neighboring provinces experienced varying patterns of excess mortality during the first wave of COVID-19. The bottom two panels of Figure 6 show excess mortality curves for Lodi and Varese, two provinces that both share a border with Milan. Excess mortality in Lodi peaked on the week of March 11, with an SMR of over 6 times the baseline mortality rate; meanwhile, excess mortality in Varese peaked a month later on the week of 8 April, with mortality for that week peaking at over double the baseline rate.

## 4 Discussion

In this study, we explored whether an application of small-area methods to excess mortality analysis could identify previously unreported trends in the pattern of excess mortality across Italy during the first wave of the COVID-19 pandemic. Our findings suggest that a small-area model yields estimates of excess mortality that are consistent with alternative calculation strategies at the national level, while offering new insights into the uneven distribution of excess mortality by age group, sex, province, and week across the country. Excess mortality estimates generated by this model suggest that a disproportionate majority of excess deaths occurred in adults age 60 and older, due to both the higher level of baseline mortality in these age groups and higher elevation of mortality above baseline during the first three months of the COVID-19 epidemic. This analysis also revealed a highly uneven spatial distribution of excess mortality: half of excess deaths were contained within just seven of Italy’s 107 provinces, accounting for less than 16% of the population. While the general nationwide pattern of excess mortality reflected the timing and geographical concentration of registered COVID-19 deaths, regional analyses obscure meaningfully different excess mortality trends across neighboring provinces within a region.

This study extends regression-based methods for estimating age-structured baseline mortality by incorporating location, year, and week structured random effects within a Bayesian hierarchical framework^7,19^. This method was found to substantially outperform a simpler approach, in which average death counts across past years are used as the baseline, for predicting weekly excess mortality. It appears that a structured space-time approach stabilizes stochastic variation across relatively small death counts by province and week, producing a smoother mortality risk surface while still accounting for meaningful trends captured by covariates. Because this approach structures uncertainty in a way that allows for principled aggregation, the results can indicate high-risk subgroups by age or location, and identify local variation in excess mortality that might be masked at a less detailed level, without overstating the confidence of findings for individual subpopulations.

This method for measuring excess mortality also has several limitations that should be noted. Because the process for estimating baseline deaths is more complicated and requires additional inputs compared to a simpler averaging method, it is less accessible to a wide range of users. The model for estimating baseline mortality assumes the same relationship between each covariate and mortality across age groups. In reality, some covariates may have a differential effect by age—for example, temperature may have more of an impact on mortality in older age groups due to the greater prevalence of risk factors that inhibit the body’s thermoregulatory response^20,21^. This limitation is partly addressed by the separate harmonic seasonality fits for each age group. This study is also limited to the set of covariates which can be estimated by province and year: other covariates that may be predictive of all-cause mortality, such as the prevalence of environmental and occupational risk factors, were excluded due to lack of availability at the province level. While the population groupings reported in this study could be divided into even more granular units, any small-area investigation must protect the privacy rights of individuals^22^. Finally, as described in the Introduction, findings from excess mortality analyses must be carefully interpreted due to the many possible sources for changing mortality which are not accounted for in the modeling strategy.

Additional mechanisms for public health monitoring are needed to catch future resurgences of COVID-19 and future epidemics. In the context of high-income countries such as Italy, where high-quality mortality data has been rapidly prepared and cleaned for public use, this approach to small-area excess mortality analysis could be employed as a routine surveillance tool, allowing health officials to identify high-mortality subgroups in a population and to introduce intervention measures in a timely manner. This approach could also be applied in many countries across Latin America which maintain high-quality mortality registration systems^23^, but where increases in COVID-19 mortality early in the pandemic have outstripped diagnostic capacity in some countries^24^. Combining this excess mortality data with cause-of-death information by province would also reveal new insights about the local drivers of excess mortality. We hope that this study provides a new avenue to convert excess mortality analysis into a tool for decision-making in public health.

## 5 Methods

### 5.1 Overview

We fitted a spatially-explicit hierarchical model with fixed effects by age group and for seven covariates, correlated age-province-year structured random effects, and harmonic curves capturing seasonal variation for each age group and province. Separate models were fit for each sex. We fit this model using mortality and population data from 1 January 2015 through 25 February 2020, then generated 1,000 predictive samples of the baseline mortality rate for each sex, age group, and province for the weeks of 26 February through 31 August 2020. For each of the 1,000 sampled draws, we compared the baseline mortality rate with the observed mortality rate to estimate a Standardized Mortality Ratio, and compared the predicted baseline deaths with observed deaths to estimate excess deaths. The code used to produce this model can be accessed online at https://github.com/njhenry/covidemr.

### 5.2 Data

Mortality and population data were downloaded from Istat, the Italian National Institute of Statistics. As of 22 October 2020, complete mortality data covering all provinces and municipalities of Italy over the time period 1 January 2015 through 31 August 2020 was available for download from Istat^25^. The number of deaths over this time period were recorded by year, month, day, Italian municipality, sex, and five-year age group. For the purposes of analysis, these observations were aggregated by sex, Italian province, age group, and week of the year. The five age groups used in this analysis were 0-59 years, 60-69 years, 70-79 years, 80-89 years, and 90+ years of age. These age groups were chosen based on the prior knowledge that the large majority of both all-cause mortality and registered COVID-19 deaths occurred among adults aged 60 and above. Weeks of the year were assigned based on the numeric day of the year, where January 1st of each year was assigned as the first day of the first week. The 365th and 366th days of the year were assigned to week 52, with the hierarchical model adjusting for observed weeks with more than seven days.

Population data by sex, age, and province for the years 2015 through 2020 was downloaded from the Istat web data portal^26^. Population counts were aggregated by sex, province, year, and the five age groups listed above.

We downloaded or extracted data for each of seven covariates, listed below in Table 2. After extraction, all covariates were normalized and rescaled to have a mean of zero and a standard deviation of 1 across all data observations.

**Table 2:** Covariates used to estimate baseline mortality by age group, sex, province, and week across Italy from January 2015 through August 2020. The source and space-time resolution of each covariate is listed.

| Covariate | Source | Varies Over |
| --- | --- | --- |
| Total fertility rate | Istat | Province, Year |
| First quarter unemployment | Istat | Province, Sex, Year |
| Proportion of eligible households receiving at-home social services | Istat | Province, Year |
| Proportion of households with taxable annual income below 10,000 Euros | Istat | Province, Year |
| Average driving time to the nearest health facility | Malaria Atlas Project | Province |
| Average elevation of residence | US Geological Survey | Province |
| Temperature | MeteoStat | Province, Year, Week |

### 5.3 Space-time model

To construct a mortality baseline for the months of March through August 2020 that incorporated multiple sources of uncertainty, we fit a small area model with age and covariate fixed effects, correlated province-year-age errors, and harmonic terms to capture seasonality within each age grouping and province. Because the age structure of mortality might differ by sex in Italy, two models were fit for males and females. For a particular sex, the number of deaths in a given province *p*, age group *a*, year *t*, and week of the year *w* was assumed to follow a Poisson distribution:

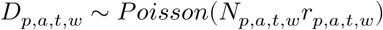

In the formulation above, *D* is the number of observed deaths, *N* is the population, and *r* is the underlying mortality rate per person-week. The quantity *r* is then fit in log space to a space-time surface which varies by province, age, year, and week:

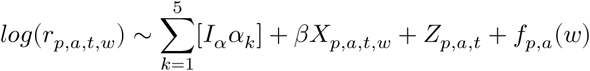

The first three terms on the right-hand side of this equation capture age and covariate fixed effects, corresponding to a discrete-time proportional hazards model where the baseline hazard varies by age group^27,28^. In this specification, *α*_*k*_ is the weekly baseline hazard for each of the five age groups, while *I*_α_ is a boolean variable that is 1 when the age group index of an observation is equal to *k* and zero otherwise. Fixed effects for the covariate design matrix *X*_*p,a,t,w*_ are denoted by *β*, a vector of length seven. Together, these terms correspond with a multivariate regression approach to estimating baseline mortality^19^.

The term *Z*_*p,a,t*,_ is a structured random effect that accounts for residual variation across provinces, age groups, and years that is not captured by the age or covariate fixed effects. *Z* is structured as a Gaussian process with mean zero and covariance matrix *K*, where *K* is a separable process across the dimensions of space, age, and time: *K* = Σ_*p*_⊗ Σ_*a*_⊗ Σ_*t*_. The spatial covariance structure Σ_*p*_ corresponds to a conditional autoregressive (CAR) process in space^29^, while the age and temporal covariance structures both correspond to discrete autoregressive processes of order 1. Separable covariance structures have been widely used in the fields of ecology and public health to construct models across space, time, and other dimensions^30,31^, and have been found to fit a wide variety of space-time covariance structures^32^.

The term *f*_*p,a*_ (*w*) refers to a set of harmonic functions that are fit to account for weekly variation in mortality not captured by covariates. A separate function is fit for each age group and province to account for the fact that seasonal variation in mortality may be driven by different factors across space and by age group. Each function is tuned to fit the parameters *A* and *B* to the following harmonics:

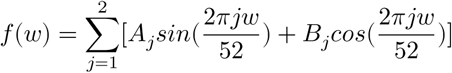

This harmonic series, which adapts principles from Fourier analysis, is the basis for a classic model for predicting seasonality in flu mortality developed by Robert Serfling^7^. In Serfling’s original formulation as well as more recent excess mortality papers, seasonality was fit using two Fourier terms^8,33^. We performed five-fold cross-validation estimate the best grouping variables and harmonic terms for seasonal curve fits. Based on the metrics of out-of-sample mean squared error and coverage, we found that the model performed best when seasonal curves were fit separately by province and age group, using two Fourier terms.

We assigned priors to all model parameters and then fit the model using the Laplace approximation for mixed-effect parameter estimation^34,35^. The model was fit in R v.4.0.3 using the package Template Model Builder v.1.7.18^34,36^.

### 5.4 Compiling and interpreting results

Using the maximum a posteriori predictions and joint precision matrix for all parameters, we generated 1,000 samples for all model parameters using a multivariate-normal approximation of the posterior predictive distribution. These parameter samples were then entered into the original model to construct 1,000 draws or “candidate maps” estimating the mortality rate across all provinces, age groups, and weeks in the study period^37^. Although the model was fit to data from 1 January 2015 through 25 February 2020, the fitted parameter fixed effects, random effects, and seasonality terms could all be applied forward to estimate 1,000 draws of predicted baseline mortality from 26 February through 31 August 2020. All subsequent calculations were performed across draws to preserve the correlation structure within draws as well as the model uncertainty across draws.

We compared the distribution of predicted mortality rates with observed mortality rates, calculated as observed deaths divided by population, to calculate 1,000 draws of standardized mortality ratios (SMRs) for each province-age-sex-year-week grouping *g* using the following formula:

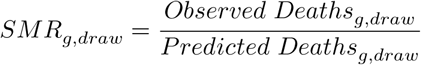

We also multiplied the predicted mortality rates by the population in each province-age-sex-year grouping to calculate predicted baseline death counts for each draw. We then calculated 1,000 draws of excess deaths for each grouping:

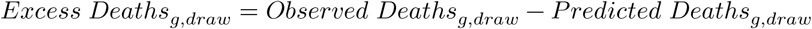

In the results section below, draws for predicted mortality, SMRs, and excess deaths are summarized using the mean and 95% uncertainty interval bounds. The 95% uncertainty interval is reported as the 2.5th percentile and 97.5th percentile of values across 1,000 draws.

### 5.5 Model validation

We used five-fold cross validation to compare predictive performance across multiple model specifications and to compare predictive performance with simpler models for calculating excess mortality. Each fold was created by fitting the model without data from the weeks in March through December for each of the years 2015 through 2019, then comparing predicted values for the held out weeks with the observed values. This holdout strategy mirrors the process we hope to capture in the months of March through August 2020 in the counterfactual where COVID-19 did not change the pattern of mortality across Italy.

Because the expected number of deaths in a given province-age-sex-year-week groupings can be very low, particularly in lower age groups, we aggregated all out-of-sample observations across four-week intervals while preserving the other groupings. We then calculated the difference between the out-of-sample recorded deaths and the modeled mortality, and calculated summary metrics: root mean squared error, coverage of the 95% uncertainty intervals, and relative squared error when compared to a simpler model that uses the average mortality rate across all other years.

We found that the out-of-sample root mean squared error for the best-performing model was 2.32E-5, compared to an average weekly mortality rate of 2.05E-4 across all age groups, suggesting a reasonably good fit for the model’s mean estimates. The out-of-sample relative squared error was 0.330 compared to the simple method of averaging weekly values across other years, suggesting that this predictive model substantially outperformed the simpler alternative for the years 2015-2019 even when an entire year of data was held out. The in-sample relative squared error compared to the simpler averaging method was 0.273, a much lower ratio of error, which indicates that the model provides a more flexible fit to the data than the simpler averaging strategy. The out-of-sample coverage of the 95% uncertainty interval was 99.1%, indicating that the predicted uncertainty bounds are conservative. The procedure for out-of-sample validation and results are discussed in more detail in the Supplementary Appendix.

## Data Availability

This research was conducted using publicly-available datasets described in the article and supplementary appendices.

https://github.com/njhenry/covidemr

## 6 Acknowledgments

The authors wish to thank the Italian National Institute of Statistics for their timely compilation and dissemination of complete vital registration data for Italy, 2015-2020, which enabled this investigation. AE would like to thank the Bill & Melinda Gates Foundation for funding his research (OPP1152978). CEM is funded by the United Kingdom’s Department of Health and Social Care, the Fleming Fund, the Wellcome Trust (grant number 209142/Z/17/Z), and the Bill and Melinda Gates Foundation (grant number OPP1176062). The computational aspects of this research were supported by the Wellcome Trust Core Award Grant Number 203141/Z/16/Z and the NIHR Oxford BRC. The views expressed are those of the authors and not necessarily those of the NHS, the NIHR or the Department of Health.

## 7 Author contributions

NH and CEM conceived and planned the study. NH identified, extracted, and processed the input data. NH carried out the statistical analyses with assistance and input from AE, MN, and MC. NH wrote the first draft of the manuscript with assistance from CEM and AE, and all authors contributed to subsequent revisions. All authors provided intellectual input into aspects of this study.

