## Supplementary methods for "Variation in COVID-19 excess mortality by age, sex, and province within Italy"

### Supplemental methods

#### Covariate preparation

Supplementary Table 1 lists the predictive covariates used to estimate baseline mortality over the study time period, 2015-2020.

| Covariate | Source | Resolution | Years covered |
|---|---|---|---|
| Total fertility rate | Istat[1] | Province, year | 2015-2018 |
| Proportion of targeted families receiving at-home social services | Istat[1] | Province, year | 2015-2017 |
| Unemployment, age 15 and above | Istat[1] | Province, year, sex | 2015-2019 |
| Proportion of households with taxable income under 10,000 Euros | Istat[1] | Commune, year | 2015-2018 |
| Average taxable income across all households | Istat[1] | Commune, year | 2015-2018 |
| Elevation | US Geological Survey[2] | 15 arc-second (gridded) | Synoptic |
| Travel time to nearest health facility by motor vehicle | Malaria Atlas Project[3] | 1 km x 1km (gridded) | Synoptic |
| Mean daily temperature | Meteostat[4,5] | Point location, day | 2015-2020 |

**Supplementary Table 1**: Summary of predictive covariates fitted in the model for baseline mortality

Synoptic and single-year covariates were used across all years, while time-varying covariates not available for the final years of the time series were projected forwards by replicating values from the final observed year of data. Covariates observed at a more detailed spatial resolution than provinces were aggregated to the province level using a population-weighted mean across units within a province.

*Weekly temperature covariate*

Point estimates of daily temperature were downloaded for the three most populous pixels in each province using the Meteostat API. Because temperature was not recorded for all time periods and locations, the following process was used to fill temperatures across all provinces and weeks in the time series:

1. Find average weekly temperatures across all observed days by year, week, and observed location.
2. Interpolate by week. In cases where 3 consecutive weeks or fewer are missing between observed temperature values in a location, estimate missing temperatures by interpolating between the nearest observed weeks

3. Aggregate by province. For each province, year, and week of the year, average available observation across the three sampled locations.
4. In rare instances where all observations for a province were missing for a given province, year, and week, temperature was filled with recorded observations from neighboring provinces with a similar elevation and level of solar exposure.

Additional covariate preparation information and analytical code can be accessed online at https://github.com/njhenry/covidemr.

#### Model validation

See the Methods section for an overview of the model validation process. We compared predictive validity metrics across six model specifications, denoted A through G. Each model specification was fit under two sets of conditions. For in-sample testing, we fit each model type using the full baseline mortality dataset from January 2015 through February 2020, and then compared estimates of underlying mortality rates with the observed data. However, the resulting in-sample metrics for goodness of fit can mask model overfitting, allowing overly flexible models to follow spurious local trends in the data.

To generate more robust estimates of model predictive validity, we also conducted out-of-sample testing that mimicked the data generation process for all-cause mortality data under a hypothetical baseline where no mortality shock occurred in 2020. Five data holdouts were created, where each holdout was missing observations from March through December in one of the five baseline years 2015-2019. We then fit each model specification based on all remaining observations, and then generated predictive validity metrics by comparing mortality rate estimates and observed data from only the held-out weeks.

As described in the Methods section, when the number of observed outcomes is low, it may be advantageous to aggregate the results across multiple dimensions of interest before generating predictive validity estimates. For this analysis, all predictive validity metrics were generated for individual observations (disaggregated by province, age group, year, and week) as well as grouped mortality rates across each province-year.

Supplementary Tables 2 and 3 report in-sample predictive validity metrics for the six model specifications when compared by province-age-year-week and by province-year groupings, respectively. Supplementary Tables 4 and 5 report out-of-sample predictive validity metrics for the six model specifications when compared by province-age-year-week and by province-year groupings, respectively. For all comparisons, root mean squared error, squared error relative to a simple mean of all observations, and squared error relative to a simpler model that uses the mean observed mortality rate for each age group were generated. We also report empirical coverage of the 50%, 80%, 90%,

95%, and 99% uncertainty intervals of each model type. Because all models are run by sex, sex-specific results as well as averages across male and female models are reported.

| Model Specification | Number of Fourier terms | Seasonality fit separately by: | Sex | RMSE | RSE | RSE compared to age-specific mean | 50% UI | 80% UI | 90% UI | 95% UI | 99% UI |
|---|---|---|---|---|---|---|---|---|---|---|---|
| A | 3 | Age, province | Male | 9.774E-04 | 0.1571 | 0.8612 | 0.6174 | 0.8629 | 0.9351 | 0.9686 | 0.9929 |
| A | 3 | Age, province | Female | 5.723E-04 | 0.0913 | 0.8667 | 0.6335 | 0.8683 | 0.9372 | 0.9692 | 0.9924 |
| A | 3 | Age, province | (Average) | 7.748E-04 | 0.1242 | 0.8639 | 0.6255 | 0.8656 | 0.9362 | 0.9689 | 0.9927 |
| B | 2 | Age, province | Male | 9.806E-04 | 0.1581 | 0.8667 | 0.6182 | 0.8642 | 0.9365 | 0.9696 | 0.9932 |
| B | 2 | Age, province | Female | 5.744E-04 | 0.0920 | 0.8731 | 0.6344 | 0.8699 | 0.9392 | 0.9701 | 0.9928 |
| B | 2 | Age, province | (Average) | 7.775E-04 | 0.1250 | 0.8699 | 0.6263 | 0.8671 | 0.9379 | 0.9699 | 0.9930 |
| C | 1 | Age, province | Male | 9.908E-04 | 0.1614 | 0.8849 | 0.6080 | 0.8546 | 0.9281 | 0.9639 | 0.9910 |
| C | 1 | Age, province | Female | 5.866E-04 | 0.0960 | 0.9107 | 0.6223 | 0.8584 | 0.9295 | 0.9635 | 0.9895 |
| C | 1 | Age, province | (Average) | 7.887E-04 | 0.1287 | 0.8978 | 0.6151 | 0.8565 | 0.9288 | 0.9637 | 0.9903 |
| D | 3 | Province | Male | 9.942E-04 | 0.1625 | 0.8910 | 0.6085 | 0.8545 | 0.9298 | 0.9652 | 0.9917 |
| D | 3 | Province | Female | 5.850E-04 | 0.0954 | 0.9056 | 0.6263 | 0.8639 | 0.9335 | 0.9668 | 0.9916 |
| D | 3 | Province | (Average) | 7.896E-04 | 0.1290 | 0.8983 | 0.6174 | 0.8592 | 0.9317 | 0.9660 | 0.9917 |
| E | 2 | Province | Male | 9.945E-04 | 0.1626 | 0.8915 | 0.6067 | 0.8531 | 0.9283 | 0.9640 | 0.9914 |
| E | 2 | Province | Female | 5.820E-04 | 0.0944 | 0.8964 | 0.6300 | 0.8683 | 0.9375 | 0.9694 | 0.9927 |
| E | 2 | Province | (Average) | 7.882E-04 | 0.1285 | 0.8939 | 0.6184 | 0.8607 | 0.9329 | 0.9667 | 0.9921 |
| F | 1 | Province | Male | 9.997E-04 | 0.1643 | 0.9010 | 0.6028 | 0.8498 | 0.9250 | 0.9617 | 0.9906 |
| F | 1 | Province | Female | 5.945E-04 | 0.0985 | 0.9353 | 0.6246 | 0.8624 | 0.9327 | 0.9662 | 0.9913 |
| F | 1 | Province | (Average) | 7.971E-04 | 0.1314 | 0.9181 | 0.6137 | 0.8561 | 0.9288 | 0.9639 | 0.9909 |
| G | (No Fourier term) | N/A | Male | 1.005E-03 | 0.1661 | 0.9105 | 0.5995 | 0.8479 | 0.9242 | 0.9610 | 0.9903 |
| G | (No Fourier term) | N/A | Female | 6.021E-04 | 0.1011 | 0.9596 | 0.6151 | 0.8526 | 0.9264 | 0.9616 | 0.9890 |
| G | (No Fourier term) | N/A | (Average) | 8.036E-04 | 0.1336 | 0.9350 | 0.6073 | 0.8502 | 0.9253 | 0.9613 | 0.9897 |

**Supplementary Table 2**: In-sample predictive validity metrics comparing model estimates to individual data observations. RMSE: root mean squared error. RSE: relative squared error.

| Model Specification | Number of Fourier terms | Seasonality fit separately by: | Sex | RMSE | RSE | 50% UI | 80% UI | 90% UI | 95% UI | 99% UI |
|---|---|---|---|---|---|---|---|---|---|---|
| A | 3 | Age, province | Male | 2.299E-05 | 0.2855 | 0.6426 | 0.9071 | 0.9577 | 0.9788 | 0.9943 |
| A | 3 | Age, province | Female | 2.547E-05 | 0.2729 | 0.6242 | 0.8929 | 0.9482 | 0.9725 | 0.9897 |
| A | 3 | Age, province | (Average) | 2.423E-05 | 0.2792 | 0.6334 | 0.9000 | 0.9530 | 0.9757 | 0.9920 |
| B | 2 | Age, province | Male | 2.268E-05 | 0.2778 | 0.6730 | 0.9210 | 0.9654 | 0.9831 | 0.9948 |
| B | 2 | Age, province | Female | 2.525E-05 | 0.2683 | 0.6532 | 0.9072 | 0.9590 | 0.9768 | 0.9920 |
| B | 2 | Age, province | (Average) | 2.396E-05 | 0.2730 | 0.6631 | 0.9141 | 0.9622 | 0.9800 | 0.9934 |
| C | 1 | Age, province | Male | 2.453E-05 | 0.3250 | 0.6064 | 0.8722 | 0.9363 | 0.9618 | 0.9848 |
| C | 1 | Age, province | Female | 2.768E-05 | 0.3225 | 0.5764 | 0.8487 | 0.9222 | 0.9527 | 0.9791 |
| C | 1 | Age, province | (Average) | 2.611E-05 | 0.3237 | 0.5914 | 0.8604 | 0.9292 | 0.9572 | 0.9819 |
| D | 3 | Province | Male | 2.332E-05 | 0.2937 | 0.6245 | 0.8980 | 0.9548 | 0.9780 | 0.9943 |
| D | 3 | Province | Female | 2.659E-05 | 0.2974 | 0.6203 | 0.8888 | 0.9501 | 0.9735 | 0.9916 |
| D | 3 | Province | (Average) | 2.495E-05 | 0.2956 | 0.6224 | 0.8934 | 0.9524 | 0.9757 | 0.9930 |
| E | 2 | Province | Male | 2.364E-05 | 0.3019 | 0.6117 | 0.8888 | 0.9489 | 0.9743 | 0.9925 |
| E | 2 | Province | Female | 2.584E-05 | 0.2809 | 0.6534 | 0.9131 | 0.9642 | 0.9813 | 0.9958 |
| E | 2 | Province | (Average) | 2.474E-05 | 0.2914 | 0.6325 | 0.9010 | 0.9565 | 0.9778 | 0.9941 |
| F | 1 | Province | Male | 2.443E-05 | 0.3223 | 0.5934 | 0.8625 | 0.9308 | 0.9626 | 0.9865 |
| F | 1 | Province | Female | 2.765E-05 | 0.3218 | 0.6027 | 0.8805 | 0.9463 | 0.9714 | 0.9884 |
| F | 1 | Province | (Average) | 2.604E-05 | 0.3220 | 0.5981 | 0.8715 | 0.9386 | 0.9670 | 0.9874 |
| G | (No Fourier term) | N/A | Male | 2.491E-05 | 0.3351 | 0.5690 | 0.8431 | 0.9233 | 0.9589 | 0.9854 |
| G | (No Fourier term) | N/A | Female | 2.862E-05 | 0.3446 | 0.5340 | 0.8234 | 0.9137 | 0.9491 | 0.9808 |
| G | (No Fourier term) | N/A | (Average) | 2.676E-05 | 0.3398 | 0.5515 | 0.8332 | 0.9185 | 0.9540 | 0.9831 |

**Supplementary Table 3**: In-sample predictive validity metrics comparing model estimates to observed data by province-year grouping. RMSE: root mean squared error. RSE: relative squared error.

| Model Specification | Number of Fourier terms | Seasonality fit separately by: | Sex | RMSE | RSE | RSE compared to age-specific mean | 50% UI | 80% UI | 90% UI | 95% UI | 99% UI |
|---|---|---|---|---|---|---|---|---|---|---|---|
| A | 3 | Age, province | Male | 9.756E-04 | 0.1727 | 0.9154 | 0.6172 | 0.8643 | 0.9364 | 0.9688 | 0.9931 |
| A | 3 | Age, province | Female | 5.683E-04 | 0.1004 | 0.9322 | 0.6370 | 0.8733 | 0.9409 | 0.9713 | 0.9937 |
| A | 3 | Age, province | (Average) | 7.720E-04 | 0.1365 | 0.9238 | 0.6271 | 0.8688 | 0.9387 | 0.9700 | 0.9934 |
| B | 2 | Age, province | Male | 9.710E-04 | 0.1710 | 0.9067 | 0.6257 | 0.8711 | 0.9406 | 0.9715 | 0.9942 |
| B | 2 | Age, province | Female | 5.660E-04 | 0.0996 | 0.9245 | 0.6441 | 0.8791 | 0.9458 | 0.9741 | 0.9940 |
| B | 2 | Age, province | (Average) | 7.685E-04 | 0.1353 | 0.9156 | 0.6349 | 0.8751 | 0.9432 | 0.9728 | 0.9941 |
| C | 1 | Age, province | Male | 9.723E-04 | 0.1715 | 0.9092 | 0.6195 | 0.8657 | 0.9370 | 0.9694 | 0.9931 |
| C | 1 | Age, province | Female | 5.733E-04 | 0.1021 | 0.9484 | 0.6402 | 0.8752 | 0.9416 | 0.9716 | 0.9931 |
| C | 1 | Age, province | (Average) | 7.728E-04 | 0.1368 | 0.9288 | 0.6299 | 0.8705 | 0.9393 | 0.9705 | 0.9931 |
| D | 3 | Province | Male | 9.695E-04 | 0.1705 | 0.9040 | 0.6320 | 0.8736 | 0.9413 | 0.9723 | 0.9939 |
| D | 3 | Province | Female | 5.824E-04 | 0.1054 | 0.9787 | 0.6521 | 0.8848 | 0.9479 | 0.9755 | 0.9946 |
| D | 3 | Province | (Average) | 7.759E-04 | 0.1380 | 0.9414 | 0.6420 | 0.8792 | 0.9446 | 0.9739 | 0.9942 |
| E | 2 | Province | Male | 9.645E-04 | 0.1688 | 0.8946 | 0.6269 | 0.8708 | 0.9404 | 0.9709 | 0.9936 |
| E | 2 | Province | Female | 5.665E-04 | 0.0998 | 0.9262 | 0.6506 | 0.8830 | 0.9467 | 0.9742 | 0.9944 |
| E | 2 | Province | (Average) | 7.655E-04 | 0.1343 | 0.9104 | 0.6388 | 0.8769 | 0.9436 | 0.9725 | 0.9940 |
| F | 1 | Province | Male | 9.745E-04 | 0.1723 | 0.9134 | 0.6266 | 0.8700 | 0.9391 | 0.9705 | 0.9933 |
| F | 1 | Province | Female | 6.022E-04 | 0.1127 | 1.0465 | 0.6492 | 0.8815 | 0.9451 | 0.9740 | 0.9939 |
| F | 1 | Province | (Average) | 7.884E-04 | 0.1425 | 0.9799 | 0.6379 | 0.8758 | 0.9421 | 0.9723 | 0.9936 |
| G | (No Fourier term) | N/A | Male | 9.906E-04 | 0.1780 | 0.9437 | 0.6189 | 0.8652 | 0.9362 | 0.9686 | 0.9930 |
| G | (No Fourier term) | N/A | Female | 6.024E-04 | 0.1128 | 1.0471 | 0.6384 | 0.8727 | 0.9406 | 0.9709 | 0.9932 |
| G | (No Fourier term) | N/A | (Average) | 7.965E-04 | 0.1454 | 0.9954 | 0.6286 | 0.8689 | 0.9384 | 0.9697 | 0.9931 |

**Supplementary Table 4**: Out-of-sample predictive validity metrics comparing model estimates to individual data observations. RMSE: root mean squared error. RSE: relative squared error.

| Model Specification | Number of Fourier terms | Seasonality fit separately by: | Sex | RMSE | RSE | 50% UI | 80% UI | 90% UI | 95% UI | 99% UI |
|---|---|---|---|---|---|---|---|---|---|---|
| A | 3 | Age, province | Male | 2.306E-05 | 0.3643 | 0.6703 | 0.9239 | 0.9725 | 0.9874 | 0.9983 |
| A | 3 | Age, province | Female | 2.513E-05 | 0.3477 | 0.6656 | 0.9125 | 0.9648 | 0.9856 | 0.9969 |
| A | 3 | Age, province | (Average) | 2.410E-05 | 0.3560 | 0.6680 | 0.9182 | 0.9686 | 0.9865 | 0.9976 |
| B | 2 | Age, province | Male | 2.205E-05 | 0.3332 | 0.7201 | 0.9455 | 0.9798 | 0.9925 | 0.9990 |
| B | 2 | Age, province | Female | 2.433E-05 | 0.3259 | 0.6975 | 0.9325 | 0.9766 | 0.9900 | 0.9986 |
| B | 2 | Age, province | (Average) | 2.319E-05 | 0.3296 | 0.7088 | 0.9390 | 0.9782 | 0.9912 | 0.9988 |
| C | 1 | Age, province | Male | 2.298E-05 | 0.3618 | 0.6647 | 0.9188 | 0.9689 | 0.9835 | 0.9971 |
| C | 1 | Age, province | Female | 2.589E-05 | 0.3691 | 0.6503 | 0.9030 | 0.9602 | 0.9818 | 0.9947 |
| C | 1 | Age, province | (Average) | 2.444E-05 | 0.3654 | 0.6575 | 0.9109 | 0.9646 | 0.9827 | 0.9959 |
| D | 3 | Province | Male | 2.223E-05 | 0.3384 | 0.7303 | 0.9477 | 0.9832 | 0.9946 | 0.9993 |
| D | 3 | Province | Female | 2.679E-05 | 0.3953 | 0.7140 | 0.9455 | 0.9837 | 0.9934 | 0.9990 |
| D | 3 | Province | (Average) | 2.451E-05 | 0.3669 | 0.7222 | 0.9466 | 0.9834 | 0.9940 | 0.9992 |
| E | 2 | Province | Male | 2.254E-05 | 0.3479 | 0.7062 | 0.9410 | 0.9786 | 0.9922 | 0.9995 |
| E | 2 | Province | Female | 2.517E-05 | 0.3488 | 0.7001 | 0.9409 | 0.9815 | 0.9922 | 0.9992 |
| E | 2 | Province | (Average) | 2.385E-05 | 0.3484 | 0.7031 | 0.9410 | 0.9800 | 0.9922 | 0.9993 |
| F | 1 | Province | Male | 2.320E-05 | 0.3687 | 0.6892 | 0.9283 | 0.9715 | 0.9866 | 0.9971 |
| F | 1 | Province | Female | 2.876E-05 | 0.4553 | 0.6754 | 0.9201 | 0.9698 | 0.9873 | 0.9976 |
| F | 1 | Province | (Average) | 2.598E-05 | 0.4120 | 0.6823 | 0.9242 | 0.9706 | 0.9869 | 0.9974 |
| G | (No Fourier term) | N/A | Male | 2.605E-05 | 0.4647 | 0.6112 | 0.8916 | 0.9545 | 0.9825 | 0.9952 |
| G | (No Fourier term) | N/A | Female | 3.044E-05 | 0.5101 | 0.5934 | 0.8748 | 0.9456 | 0.9755 | 0.9946 |
| G | (No Fourier term) | N/A | (Average) | 2.824E-05 | 0.4874 | 0.6023 | 0.8832 | 0.9500 | 0.9790 | 0.9949 |

**Supplementary Table 5**: Out-of-sample predictive validity metrics comparing model estimates to observed data by province-year grouping. RMSE: root mean squared error. RSE: relative squared error.

Based on out-of-sample predictive validity metrics by province-year grouping, as summarized in Table 5, we selected specification B as the model type that was used in the final analysis. This model specification had the lowest root mean squared error across both male and female runs, with slightly conservative empirical coverage of the 95% and 99% uncertainty intervals. Out-of-sample predictive validity metrics did not strongly differentiate between the model specifications that included any seasonality terms. However, the model specification that included no seasonality term performed worst in terms of both in-sample and out-of-sample predictive validity.

#### Excess mortality time series estimation

When calculating a time series of excess mortality among small population groups, the cumulative effect of past mortality on the base population must be taken into account. This problem is most clearly illustrated under artificial conditions of a very high mortality rate. Imagine a population of 1,000 individuals that, under normal conditions, experiences a baseline mortality rate of 100 deaths per 1,000 person-weeks and predictably gains 100 new members at the end of each week. Under normal conditions, 100 individuals from the population would die and 100 new individuals would be added each week, resulting in a stable population week-to-week. Now, imagine an event that causes the mortality rate to increase to 900 deaths per 1,000 person-weeks. In the first week under these conditions, 900 individuals from the base population of 1,000 would die and 100 would be added, leading to a population of 200 entering the second week. In the second week, 180 of 200 individuals in the population would die. Although the standardized mortality ratio of this event compared to baseline is 9 across both weeks (900/100 and 180/20), an analysis that mistakenly assumed a starting population of 1,000 in the second week would dramatically underestimate this ratio as 1.8 (180/100).

This analysis took into account the effects of excess mortality on subsequent population denominators throughout the study weeks beginning on February 26, 2020. Population denominators listed by Istat for January 1, 2020 were used as the starting denominators for this time series. For each predictive posterior draw and week, calculated excess deaths were subtracted from the base population used as the denominator in the subsequent week. The cumulative effect of this correction is to avoid understating the toll of excess mortality over the time period due to a reduction in the base population.
